# Chemical/technical risk analysis of a new multi-directional nebulizer (MDN) and its clinical implementation for the safe administration of Pressurized Intraperitoneal Aerosol Chemotherapy (MDN-PIPAC)

**DOI:** 10.1101/2023.12.01.23297348

**Authors:** Fabian Kockelmann, Daniel Göhler, Sarah Barbey, Amayu Wakoya Gena, Hayder Alsaad, Conrad Voelker, Jürgen Zieren, Mehdi Ouaissi, Urs Giger-Pabst

## Abstract

**Background/Aim:** To test the chemoresistance of a multi-directional nebulizer (MDN) and to establish and implement a perioperative clinical safety concept for its clinical use to deliver pressurised intraperitoneal aerosol chemotherapy (MDN-PIPAC).

**Study design:** Ex-vivo nebulization of cytostatic drugs with the MDN device to assess chemoresistance/toxicological risks. Establishment of a perioperative safety concept for the clinical administration of MDN-PIPAC by ex- and in-vivo porcine simulation studies. Unicentric case series of 30 MDN-PIPACs in 21 patients with peritoneal surface malignancies (PSM). Endpoints were intraoperative adverse events and perioperative complications (Clavien-Dindo).

**Results:** Toxicological studies/risk assessment confirm the safety of administering PIPAC with the MDN. The horizontal nozzles must protrude at least 7 mm beyond the most distal end of the trocar tip and lateral tilting should be prevented by fixation of the device in a single-arm holder. A total of 21 patients (male/female ratio: 2:1) with a mean age of 62 (range: 38-86) years underwent 30 consecutive MDN-PIPACs for peritoneal surface malignancies of different origin. ECOG 0 and 1 were seen in five and 16 patients, respectively. Thirteen, seven, and one patient underwent one, two, and three MDN-PIPACs, respectively. Two patients received only one cycle of MDN-PIPAC because they were considered candidates for cytoreductive surgery and heated intraperitoneal chemotherapy. There were no intraoperative technical/medical problems observed. Four patients suffered from postoperative grade I complications.

**Conclusions:** Compounds leached during chemotherapy nebulization with the MDN are toxicologically safe. MDN-PIPAC administrations are safe and the postoperative course is comparable to that of the conventional PIPAC nebulizer.

**MINI ABSTRACT:** Toxicological analyses confirm that doxorubicin, cisplatin, and oxaliplatin nebulized with the MDN do not release leachables that pose a toxicological risk to patients. Through technical risk analysis and PIPAC simulations, a safety concept for the administration of MDN-PIPAC was established. No technical/medical intraoperative adverse events were observed. The perioperative course was comparable to that of the conventional axial single-direction nebulizer (SDN) PIPAC.

## 1.0 INTRODUCTION

For more than a decade, pressurised intraperitoneal aerosol chemotherapy (PIPAC) has been used for the treatment of peritoneal surface malignancies (PSM). Clinical results are encouraging, but data from prospective, randomized trials clearly demonstrating the efficacy of this treatment are still lacking [1, 2]. A common feature of all previously available nebulizers for PIPAC therapy is that they all have only one nebulizer unit (axial single-direction nebulizer (SDN)), which ejects the chemotherapeutic droplets in an axial direction. The majority (97.5 % of the volume) of the nebulized chemotherapy impacts the peritoneum directly, which is located below the nebulizer orifice [3]. It has also been suggested that the locally high drug deposition that occurs when using a SDN results in potential local toxicity that could lead to an increase in perioperative complications, particularly when additional surgical procedures such as complex adhesiolysis or bowel resection are performed at the same time as PIPAC [4 - 6]. However, more recent clinical data appear to relativise such assumptions [7].

However, the PIPAC nebulizer technology has been further developed to optimize the spatial distribution of the drug. A new and innovative concept is multi-directional drug aerosolization. Recent data obtained with a multi-directional drug nebulizer prototype reported an improved spatial drug distribution pattern in an ex-vivo PIPAC model compared to standard SDN technology [8].

A first multi-directional nebulizer (MDN) for the clinical off-label application of PIPAC (MDN-PIPAC) is now commercially available. In addition to the conventional axial nozzle, three additional horizontal nozzles are integrated into the nebulizer head at 120° intervals. This nozzle configuration enables nebulization of the chemotherapy over a spray angle of almost 240° - compared to 77° with a SDN [9]. Since there are still no documented preclinical or clinical applications with this device worldwide, we decided to conduct further extensive preclinical studies before the first clinical MDN-PIPAC applications.

Therefore, the objectives of this study were (i) to investigate a potential toxicological risk from substances leaking from the device when used in combination with cytotoxic drugs, and (ii) to develop an intraoperative safety concept for the administration of MDN-PIPAC using an ex-vivo PIPAC model, schlieren imaging, and in-vivo large animal CT peritoneography, and iii) to analyse a consecutive case series of 30 MDN-PIPACs to evaluate perioperative safety and feasibility in patients with peritoneal surface malignancies (PSM).

## 2.0 MATERIAL & METHODS

### 2.1 Research sites and institutes

Nebulization for cytostatic drug compatibility/toxicology studies of the MDN and the in-vivo animal experiments were carried out at the French National Research Institute for Agriculture, Food and Environment (INRAE), Centre Val de Loire Nouzilly, France. Schlieren imaging was performed at the Bauhaus-University Weimar, Weimar, Germany. Chemical characterisation and toxicological evaluation were conducted by RESCOLL, Pessac, France, and TentaConsult Pharma & Med GmbH, Münster, Germany, respectively.

### 2.2 MDN chemotherapy compatibility study

Doxorubicin, cisplatin, and oxaliplatin were purchased from Hexal AG, Holzkirchen, Germany, Accord Healthcare B.V. Utrecht, the Netherlands, and Medac GmbH, Wedel, Germany. Drug doses (doxorubicin 6 mg, cisplatin 30 mg, and oxaliplatin 120 mg per m^2^ of body surface area (BSA)) were studied according to data from phase I safety studies and clinical use [10- 12]. A patient with a height of 1.90 m and a weight of 100 kg was assumed. The BSA was calculated according to the DuBois formula [13]. Glucose 5% and NaCl 0.9% were purchased from B. Braun, Melsungen, Germany. Oxaliplatin was diluted with glucose 5% and doxorubicin and cisplatin with NaCl 0.9% to a total of 50 ml and 150 ml, respectively. The same solutions were used as blank controls. Blank control solutions were immediately filled into sterile glass bottles (ref. # FRAA-250-60, Labbox GmbH, Düsseldorf, Germany) and stored protected from light at 4°C. The chemotherapy solutions for nebulization experiments were filled into syringes (ELS 200 ml (S), MEDTRON AG, Saarbrücken, Germany), the syringe instated into the high-pressure injector head (Accutron® HP-D Vision, MEDTRON AG, Saarbrücken, Germany), vented, and then connected to the high-pressure line of the MDN (QuattroJet™, REGER Medizintechnik GmbH, Villigendorf, Germany). The shaft of the MDN was wrapped with sterile surgical compress proximal to the head of the nebulizer and then positioned vertically in a tight and stable position in the neck of a sterile 250 ml glass bottle (ref. # FRAA-250-60, Labbox GmbH, Düsseldorf, Germany). Nebulization could take place unhindered with the nebulizer head in the body of the bottle at a flow rate of 1.5 ml/s with an upper pressure limit of 300 psi. Nebulization was undertaken under a vertical laminar air flow bench. Doxorubicin and Cisplatin were nebulized one after the other but into the sample bottle. All experiments were performed in triplicates. The sample and blank control solution bottles were sealed airtight immediately after the experiments and stored protected from light at 4°C. The samples were transported to the laboratory for further processing by courier within 4 hours while refrigeration was maintained.

### 2.3 Chemical characterisation and toxicological risk assessment

MDN device (QuattroJet™, REGER Medizintechnik GmbH, Villigendorf, Germany) and chemical characterisation according to the ISO 10993-18:2020 [14]. Head-Space Gas Chromatography coupled with Mass Spectrometry (HSGC-MS) was used for detecting volatile organic compounds, Gas Chromatography coupled with Mass Spectrometry (GC-MS) for detecting semi-volatile organic compounds, Liquid Phase Chromatography□coupled to a Mass Spectrometer (LC-MS) for detecting non-volatile organic compounds, Inductively Coupled Plasma Atomic Emission Spectrometry (ICP-OES) for quantifying the inorganic elements, and Ion Chromatography (IC) for quantifying the ions. The Analytical Evaluation Threshold (AET) was applied. The AET defines the threshold below which leachable or extractable substances cannot be identified or quantified and therefore it can be assumed without further evaluation that such a substance does not pose a toxicological risk.

The parameters used for the toxicological risk assessment were set to consider the worst case. Three consecutive MDN-PIPAC administrations were assumed with no time interval in between (instead of a treatment interval of four to six weeks between each PIPAC cycle). In addition, the exposure of children with a body weight of 10 kg was calculated with the amount used for the treatment of an adult. The toxicological risk associated with leachables was assessed by collecting data and identifying critical health endpoints through literature searches and toxicological risk assessment from PubChem (HSDB), PubMed, ChemIDplus, ECHA databases, and various other online sources and databases (e.g., FDA.gov) in accordance with ISO 10993-17:2023 [15].

### 2.4 Preclinical assessment of the MDN-PIPAC safety checklist and schlieren imaging

As a first step, a team of two surgeons and one PIPAC nurse analysed whether the use of the MDN (QuattroJet™, REGER Medizintechnik GmbH, Villigendorf, Germany) could result in additional intraoperative risks or hazards for healthcare personnel. Potential risks were assessed for clinical relevance. Appropriate safety measures were then developed and integrated into our existing intraoperative PIPAC safety checklist [16]. In a second step, using this adapted intraoperative PIPAC safety checklist, the team repeatedly simulated MDN-PIPAC applications in the operating room using NaCl 0.9% (B. Braun, Melsungen, Germany) on an established ex-vivo PIPAC model [17] to test the coherence of the adapted intraoperative PIPAC safety measures and the safety checklist.

In addition, the schlieren imaging system of the Bauhaus-University Weimar (Figure 1) was used to investigate the potential for chemotherapy aerosol/CO_2_ backflow through the trocar valve system into the operating theatre environment during MDN-PIPAC. A worst-case scenario was simulated in which the head of the MDN is misplaced in the trocar shaft, resulting in nebulization in the trocar sleeve. Schlieren imaging is a flow visualization technique that allows for visualizing density gradients in fluids by capitalizing on the refraction of light when entering a medium of different density.

**Figure 1:**
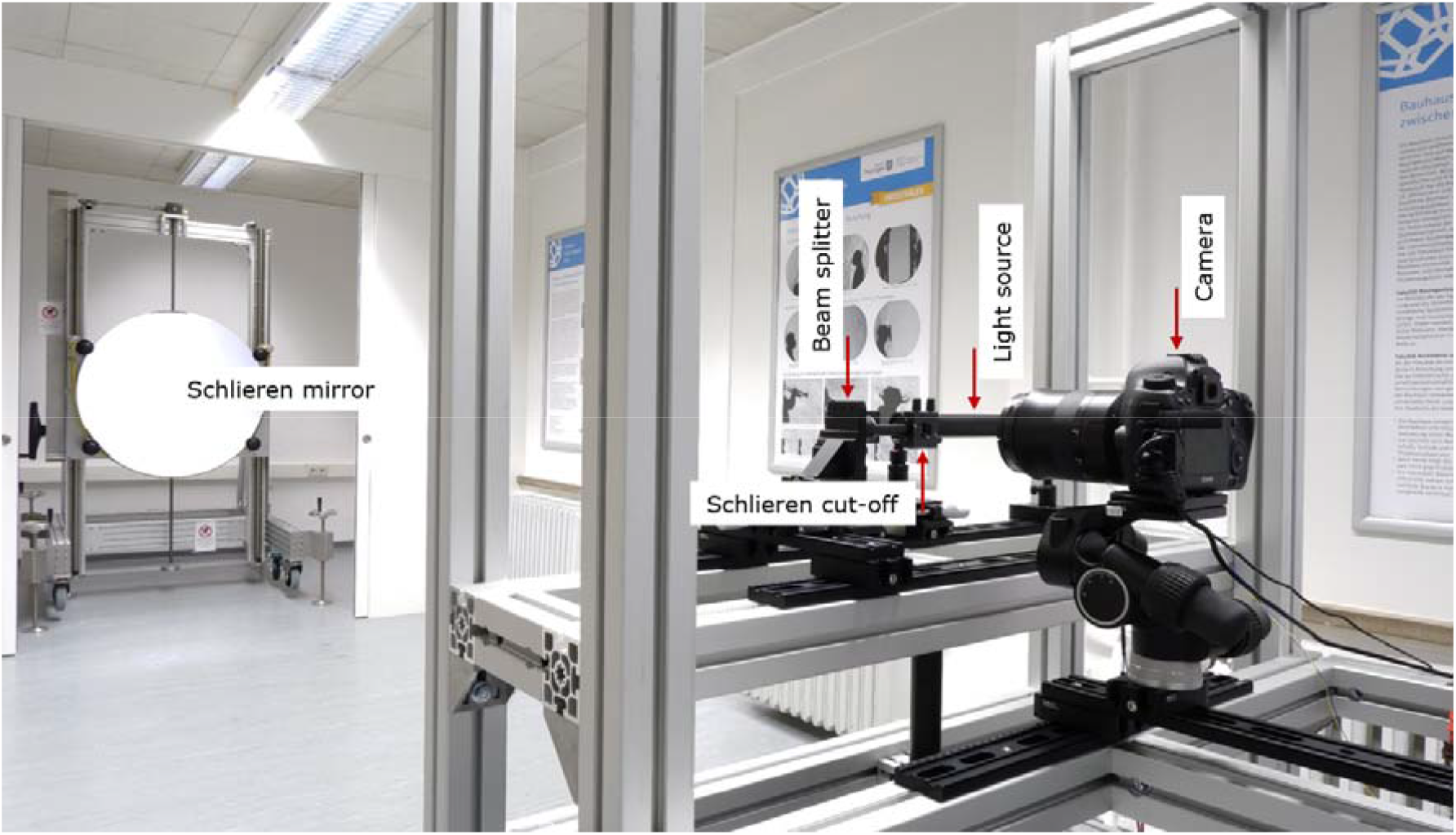
The schlieren imaging system at the Bauhaus-University Weimar

The utilized schlieren system uses a single concave spherical mirror, where the light beams from the source and the returning light rays from the mirror align coincidently, forming an on-axis single-mirror coincident schlieren system [- 20]. The system consists of five optical elements: concave spherical mirror (1 m in diameter), light source (light-emitting diode (LED) including condenser lens and pinhole), beam splitter, schlieren cut-off, and digital camera (Canon EOS 5DS R, with 50.6 megapixels image size and a 135 mm focal-length lens). The schlieren mirror has a surface accuracy of λ/9.5, where λ represents the wavelength of HeNe laser light. This high level of accuracy makes it highly sensitive, enabling the visualization of very small density gradients in fluids. In this study, the density gradient results from differences between the leaking CO_2_ and the surrounding air. To enhance visualization, the CO_2_ was heated using an insufflator (Figure 2), creating a sufficiently large density gradient for schlieren imaging. The system’s ability to visualize this density gradient was also tested prior to the investigation (Figure 5A). Throughout the investigations, the camera’s video imaging was set at 50 frames per second.

**Figure 2:**
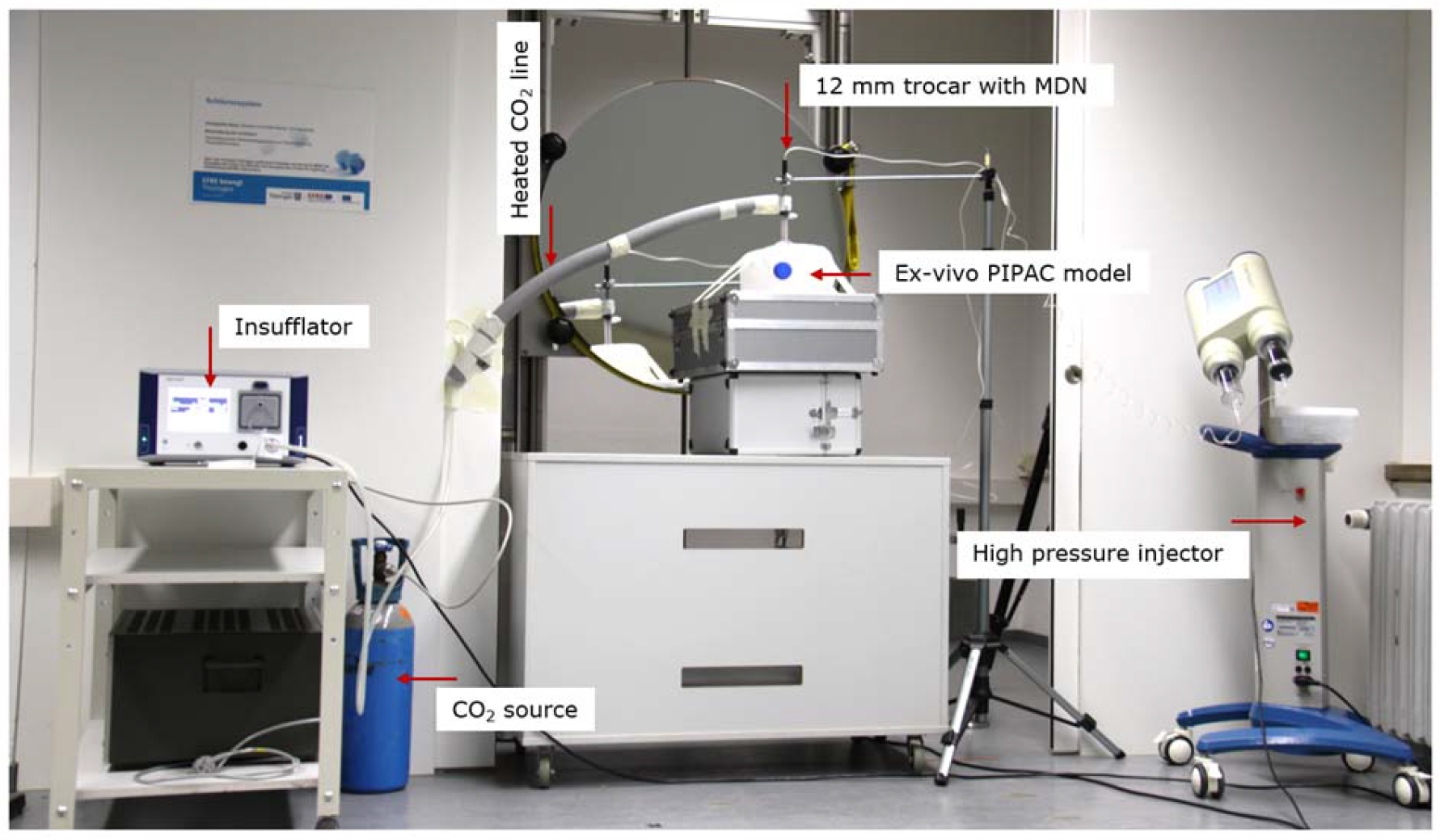
Test setup for schlieren imaging during ex-vivo MDN-PIPAC simulations

The schlieren flow visualizations investigated possible CO_2_/aerosol backflow scenarios during clinically relevant MDN-PIPAC scenarios in an ex-vivo PIPAC model with three litre volume, MDN in central, vertical position, fixed on a stand in a 12 mm balloon trocar (Kii Sleeve with advanced fixation, Applied Medical, California, USA) with lateral nebulizer nozzles that protrude 7 mm beyond the distal end of the trocar shaft. Stable capnoperitoneum of 12 or 20 mmHg was maintained throughout the whole experiments (Insufflator Flow 50, B. Braun SE, Melsungen, Germany). Injection of 50- and 150-ml distilled water at 1.5 ml flow/min (SinoPower-D, Sino Medical-Device Technology Co., Ltd, Shenzhen, China). Worst-case scenario simulation at 20 mmHg capnoperitoneum, flow rate 1.5 ml/min with incorrect positioning of the MDN head in the distal trocar shaft, so that nebulization occurred in the trocar shaft. During the flow visualizations, the ex-vivo PIPAC model was placed in front of the schlieren mirror (Figure 2).

### 2.5 In-vivo animal MDN-PIPAC and CT-peritoneography

Domestic pigs were handled and cared for according to all relevant guidelines by authorised investigators. The study (file # 40583) was approved by the French Ministry of Higher Education and Research (Ministère de l’enseignement supérieur et de la recherche). A total of three male animals weighing from 47 to 42 kg were used. Anaesthesia was induced by intramuscular injection of ketamine 20 mg/kg, xylazine 2 mg/kg, and a subcutaneous injection of atropine 0.02 mg/kg, and then completed by endotracheal intubation. The animals were maintained under anaesthesia by isoflurane 3%, intravenous sufentanil, and cisatracurium. The animals were fixed in a supine position in the CT-scanner (SOMATOM Definition AS 128, Siemens, Germany) to deliver HP/HD PIPAC described previously [12]. The MDN was placed at the centre of the abdomen in a perpendicular position so that the horizontal nozzle openings protruded approx. 7 mm beyond the end of the 12-mm trocar (Kii balloon blunt-tip access system, Applied Medical, California, USA). The orientation of these horizontal nozzle openings was chosen so that one nozzle was directed into the right and left upper abdomen and the third nozzle into the small pelvis. Another 5-mm trocar was inserted into the abdominal cavity to monitor the position of the nozzle head with a 5 mm camera and the nebulization process. CT-peritoneography was performed using a modified technique previously described in [21]. For this purpose, 100 ml Imeron® 300 (Bracco Imaging GmbH, Konstanz, Germany) was diluted in 900 ml NaCl 0.9% (B Braun, Melsungen, Germany) in a ratio of 1:10. 150 ml of this solution was then administered for HP/HP-PIPAC CT-peritoneography at a flow rate of 1.5 ml/s (Accutron CT-D Vision, MEDTRON AG, Saarbrücken, Germany). CT-peritoneography was performed immediately at the end of the HP/HD-PIPAC. After CT acquisition, the animals were immediately euthanized by intravenous injection of phenobarbital.

### 2.6 Patients’ selection criteria for MDN-PIPAC

The clinical study was performed in line with the guidelines of the Declaration of Helsinki at the Department of Surgery, Klinikum Dortmund, University Hospital of the University Witten/Herdecke, Germany. All patients gave oral and written informed consent. The approval of the Ethics Committee of the Medical Faculty of the University of Münster and the Medical Association Westphalia-Lippe in Germany (file # 2021-360-f-s) was obtained. All patients had histologically confirmed primary or metastatic peritoneal surface malignancies (PSM). Patients were selected by a multidisciplinary tumor board accredited by the German Society for General and Visceral Surgery (DGAV). All procedures were performed as previously described in detail in [10, 12] and were performed by one of the authors (J. Z., senior surgeon). The time interval between cycles was between four to six weeks. Systemic chemotherapy was discontinued one week prior and after MDN-PIPAC. Patients were informed that PIPAC treatment was not provided within the framework of evidence-based therapy guidelines. All patients gave their oral and written consent. Oxaliplatin at a dose of 120.0 mg/m^2^ body surface area (BSA) diluted in a total of 150 ml glucose 5% was administered in case of PSM of colorectal and appendiceal primary tumours. For all other tumour entities, doxorubicin 6.0 mg/m^2^ BSA diluted in 50 ml NaCl 0.9% followed by cisplatin 30.0 mg/m^2^ BSA diluted in 150 ml NaCl 0.9% was administered [- 12]. According to the manufacturer’s instructions, the MDN was operated at a liquid fluid volume flow of 1.5 ml/s with an upper pressure limit of 300 psi. Access to the peritoneal cavity was always obtained via an infraumbilical open Hasson approach. Before starting the drug nebulization, the capnoperitoneal pressure was increased from 12 to 20 mmHg and then kept constant for 30 minutes.

### 2.7 Perioperative short-term outcome and data acquisition

Retrospective data acquisition of a consecutive case series of 30 delivered MDN-PIPACs. The entire staging laparoscopy, including documentation of the Sugarbaker PCI score, the amount of ascites and the quality of chemotherapy nebulization, is monitored by video according to our standards for intraoperative documentation. All data are stored electronically in the patient’s record. Data acquisition and database management are carried out by the co-author J. Z. as part of the required quality assurance as a certified centre of the DGAV. Perioperative adverse events are graded according to the Clavien-Dindo Classification [22, 23]. Data are expressed as absolute numbers whereas continuous data are expressed as are expressed as median values followed by the range in parentheses, i.e. median (range).

## 3.0 RESULTS

### 3.1 Chemical characterisation and toxicological risk assessment

The results of the chemical analyses of nebulized chemotherapy solutions are summarized in Table 1.

**Table 1:** Toxicological analyses after nebulization of chemotherapy with the MDN. *Legend: HSGC-MS = Head-space Gas Chromatography coupled with Mass Spectrometry; GC-MS = Gas Chromatography coupled with Mass Spectrometry; LC-MS = Liquid phase chromatography..coupled to a Mass Spectrometer; ICP-OES = Inductively Coupled Plasma Atomic Emission Spectrometry; IC = Ion Chromatography;*

| Methods of Analysis | Doxorubicin + Cisplatin | Oxaliplatin |
| --- | --- | --- |
| HSGC-MS analysis for Volatile Organic Compounds | Not detected | Not detected |
| GC-MS analysis for Semi-Volatile Organic Compounds |  |  |
| - 1, 6, 11, 16-Tetraoxacycloeicosane | Not detected | 0.03 µg/ml |
| - 1, 6, 11, 16, 21-Pentaoxacyclopentacosane | Not detected | 0.04 µg/ml |
| LC-MS analysis for Non-Volatile Organic Compounds |  |  |
| - Cyclic polybutylene glycol (C20) | 0.144 µg/ml | 0.131 µg/ml |
| - Cyclic polybutylene glycol (C24) | 0.246 µg/ml | 0.123 µg/ml |
| - Cyclic polybutylene glycol (C32) | 0.083 µg/ml | 0.045 µg/ml |
| ICP-MS analysis for inorganic elements |  |  |
| - Copper | 0.049 µg/ml | Not detected |
| - Ferrum | 0.057 µg/ml | 0.011 µg/ml |
| - Manganese | 0.015 µg/ml | 0.014 µg/ml |
| IC analysis for anions |  |  |
| - Acetates | Not detected | Not detected |
| - Fluorides | Not detected | Not detected |
| - Chlorides | Not detected | Not detected |
| - Bromides | Not detected | Not detected |
| - Nitrites | Not detected | Not detected |
| - Nitrates | Not detected | Not detected |
| - Sulfates | Not detected | Not detected |
| - Phosphates | Not detected | Not detected |

The independent systematic toxicological risk assessment based on the assumption of three consecutive PIPAC sessions on the same day in a child with a body weight of 10 kg revealed no toxicological risk for the few compounds detected after nebulization.

### 3.2 Preclinical technical, safety checklist and surgical approach assessment for MDN-PIPAC

The following three relevant sub-areas were defined and perioperative measures were implemented:

#### 3.2.1 Operational parameters

According to previous research data and the manufacturer’s specifications, the MDN is operated best with a pressure of 150-250 psi with an upper safety limit pressure of 300 psi. In practice, this pressure operating window corresponds to the liquid volume flow rate of 1.2 ml/s-1.6 ml/s. [2]. Accordingly, these operating parameters were stored in the high-pressure injector as a separate MDN-PIPAC working programme. As there are no other differences in handling and technical operation compared to the conventional nebulizer used, no additional measures were necessary.

#### 3.2.2 Construction & Design

Due to the three horizontal nozzles, there is a potential risk of drug nebulization in the trocar if the horizontal nozzles are not placed sufficiently beyond the distal end of the trocar into the abdominal cavity. In the worst case, retrograde leakage of chemotherapy through the trocar valve into the operating theatre could occur and harm healthcare workers. To avoid this, the correct positioning of the nebulizer head in the trocar has been added to our standard safety checklist. The four-eye principle is used to check that the horizontal nozzles extend beyond the most distal point of the trocar. It should be noted that most trocars are bevelled at an angle of approximately 15°. The most distal end of the trocar is therefore used as the reference point (Figure 3A).

**Figure 3:**
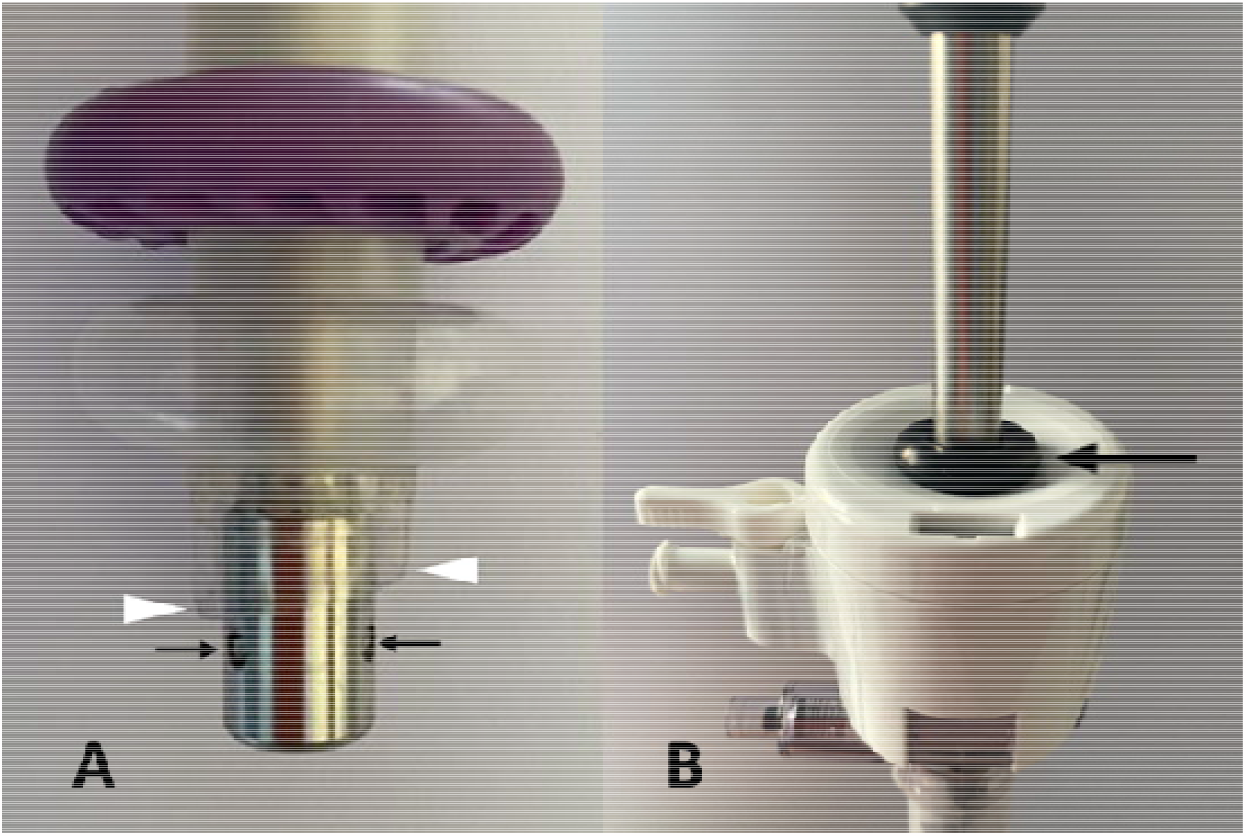
Ex-vivo evaluation of the optimal position of the MDN in the 12-mm trocar *Legend: A = MDN inserted into a 12 mm balloon trocar. The position of the MDN head must be checked visually so that the lateral nozzles do not nebulize into the trocar. The white triangles mark the distal end of the trocar. Note that the trocar is angled 15° at the distal end. The black arrows point to the horizontal nozzle openings. These should protrude at least 7 mm beyond the most distal end of the trocar (left white arrow). B = MDN inserted into a 12 mm balloon trocar. The black arrow points to the sterile rubber ring pulled over the nebulizer shaft which rests on the trocar head to prevent the nebulizer from gliding into the trocar*.

#### 3.2.3 Surgical approach

To optimize spatial aerosol distribution during MDN-PIPAC, the spraying distance between the nozzle orifice and the peritoneum should be as large as possible [2, 7]. Periumbilical, perpendicular placement of the MDN is therefore ideal, if surgically possible (Figure 4A). Additionally, we decided to orientate the MDN in a way that one horizontal spray jet pointed to the right upper, a second to the left upper abdominal quadrant, and the third towards the small pelvis (Figure 4B). In addition, we decided to routinely fix the nebulizer intraoperatively in a single-arm holder (M-TRAC, Braun, Melsungen, Germany) to further prevent tilting and axial sliding into the trocar. The central periumbilical perpendicular MDN position as well as the orientation of the horizontal nozzle orifices were integrated into our intraoperative safety checklist.

**Figure 4:**
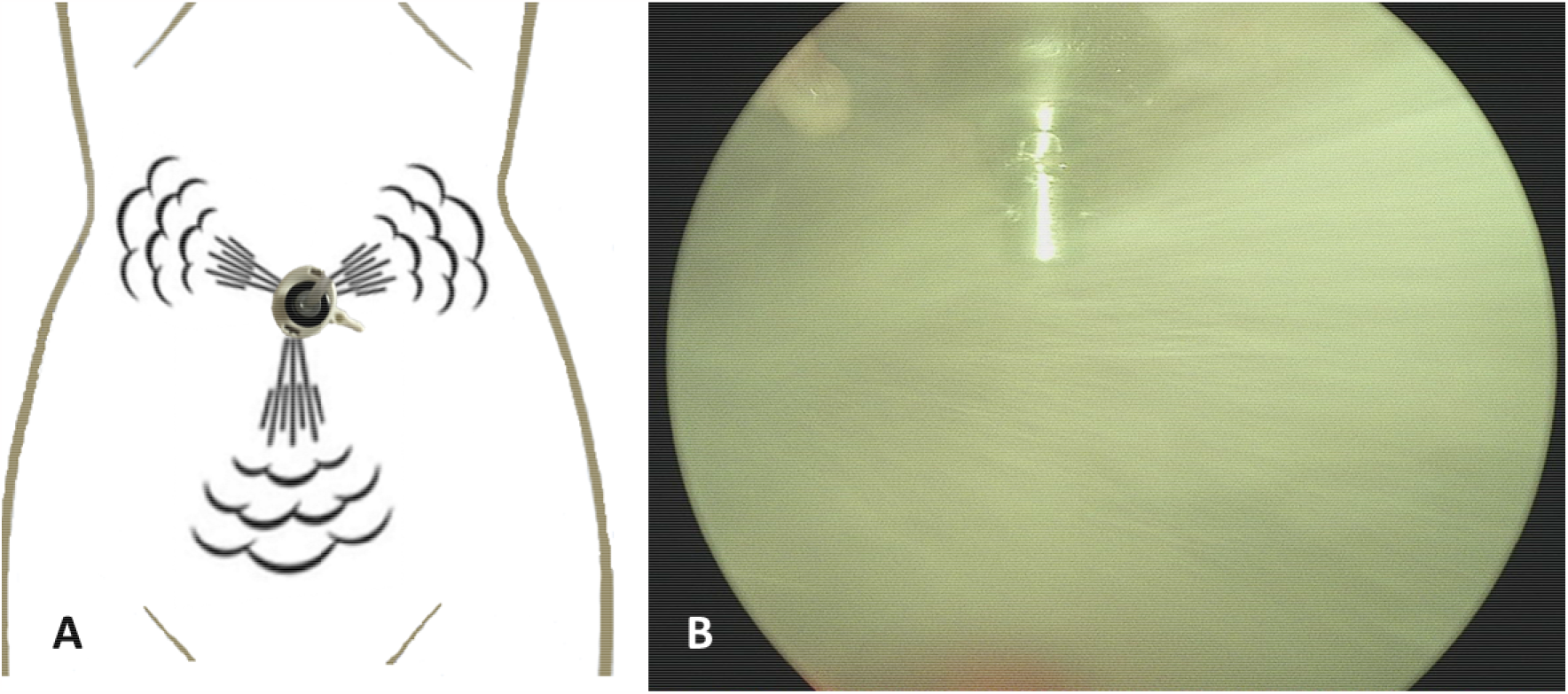
Periumbilical position of the MDN for optimized nebulization *Legend: A = Sketch of infraumbilical position of the MDN for optimized nebulization of all four nozzles (only horizontal nozzles shown); B = intraoperative view on the MDN head*

### 3.3 Ex-vivo MDN-PIPAC simulation and schlieren imaging

As the distal end of the trocar is angled, special care must be taken to ensure that the distal end of the trocar is visible intraoperatively so as not to interfere with the nebulization of the horizontal nozzles. The correct position of the MDN head has therefore been included as a separate item in our extended safety checklist. The openings of the horizontal nozzles must extend at least 7 mm beyond the distal end of the trocar to avoid interference during nebulization (Figure 3A). In addition, the vertical position of the MDN in the 10/12-mm trocar was found to cause the MDN to tend to slide inward in the trocar without additional fixation. To avoid MDN slippage into the trocar, the device is placed on the trocar head with a rubber ring (sterile, reusable trocar seal) previously pulled over the shaft of the nebulizer in such a way that the rubber ring fits tightly on the trocar head and prevents the nebuliser from slipping inside the trocar (Figure 3B). To further stabilize the position of the MDN, using a single-arm holder turned out to be essential and was therefore also included in our safety checklist. Mispositioning the MDN head with the horizontal nozzles nebulizing in the trocar shaft could cause aerosol leakage into the environment. However, multiple simulations of such a scenario showed no macroscopic detectable retrograde aerosol leakage into the environment. The further schlieren visualisations also showed no leakage of CO_2_/aerosol into the environment in any of the scenarios investigated. Even with unfavourable positioning of the MDN head with nebulization in the distal trocar shaft, there was no evidence of even a slight retrograde flow through the trocar valve. Figure 5B shows a representative schlieren image.

### 3.4 In-vivo pig MDN-PIPAC CT-peritoneography

The CT-peritoneographies of the three pigs could be performed without perioperative complications. The animals showed no cardiorespiratory problems due to the capnoperitoneal pressure of 20 mmHg. Our previously prepared MDN-PIPAC safety checklist was implemented without any problems. The subsequent CT-peritoneographies showed an excellent distribution pattern of the administered contrast agent aerosol. However, no contrast agent was visible in the ventral portions of the abdominal cavity. Figure 6 depicts representative images of pig N° 1.

**Figure 5:**
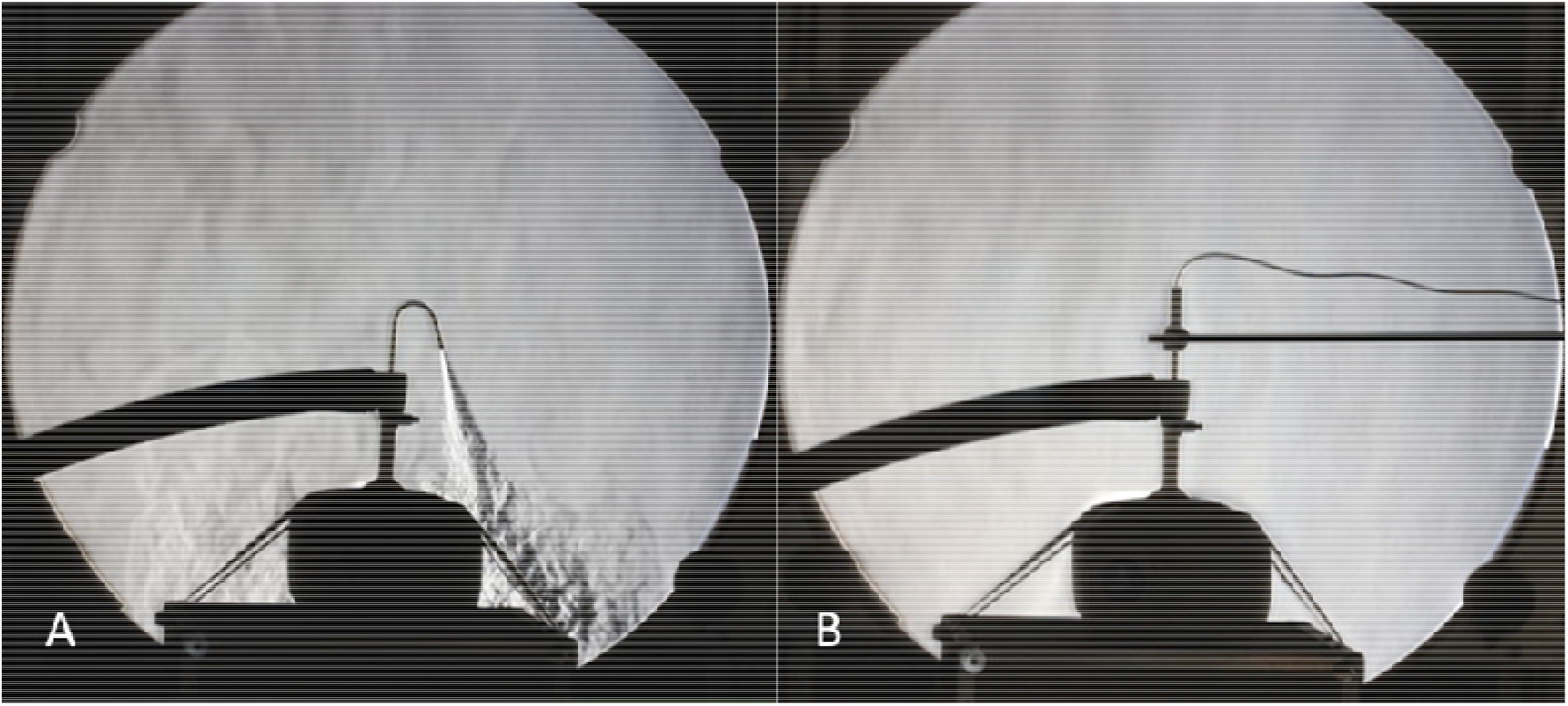
Schlieren imaging of different scenarios of MDN-PIPAC simulation *Legend: A = a test schlieren image to demonstrate the visualization of CO*_*2*_ *leakage; B = schlieren visualization of 12 mmHg injection at 1.3 ml/s with a 50 ml of distilled water*

**Figure 6:**
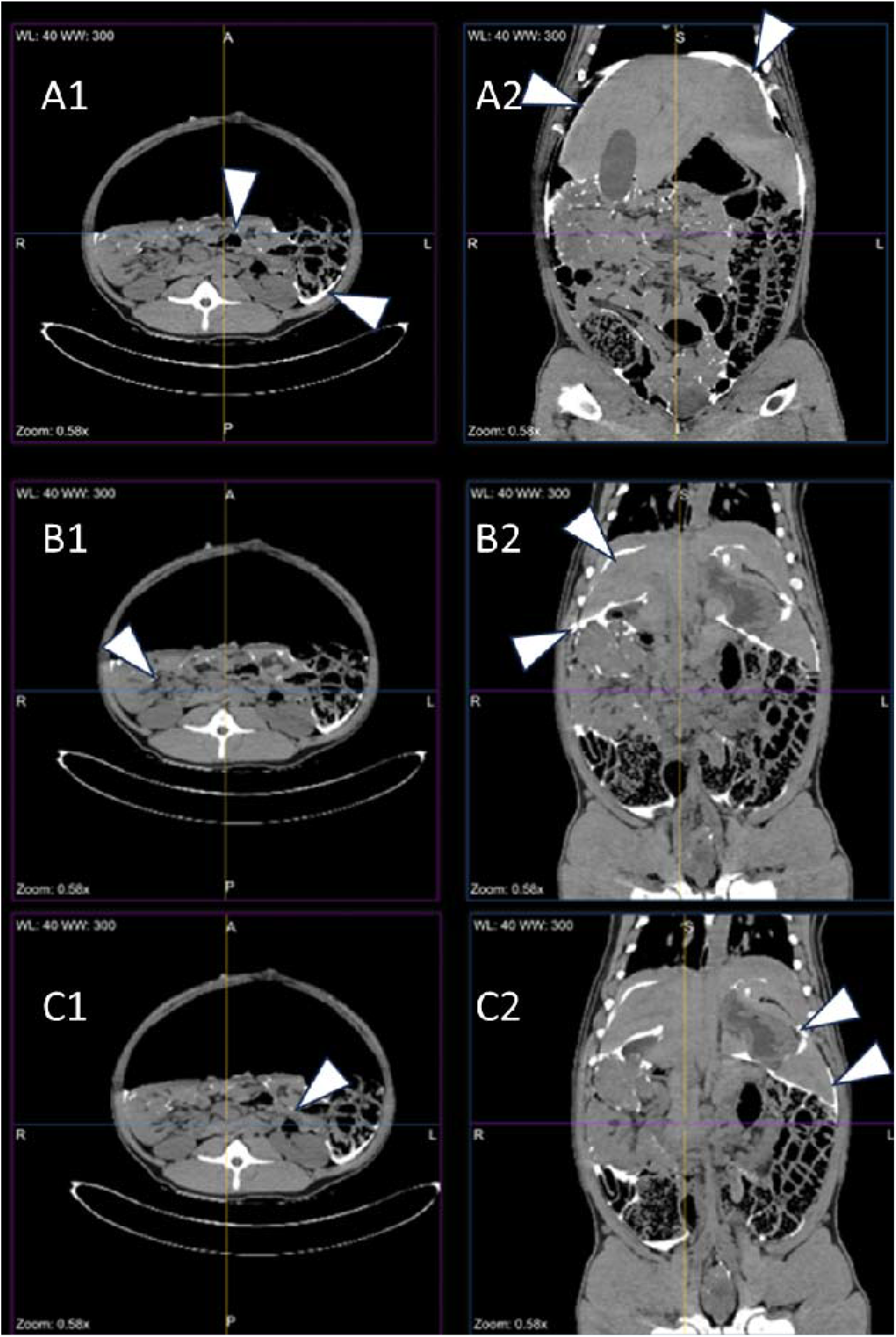
Representative MDN-PIPAC CT-peritoneography images of pig N° 1 *Legend: A1-C1 = horizontal section planes; A2-C2 = coronal section planes. The white triangles point to the contrast agent accumulation: A1-C1 = enter-enteric and paracolic left; A2 = subdiaphragmatic bilateral; B2 = homogeneous on the Glisson capsule, subhepatic and interlobar; C2 = perigastric and perilienal*

### 3.5 Clinical baseline characteristics

A total of 21 patients (male/female ratio: 2:1) with a mean age of 62 (range: 38-86) years underwent 30 consecutive MDN-PIPACs. ECOG 0 and 1 were seen in five and 16 patients, respectively. Patients had PSM of the upper gastrointestinal tract (n=6), ovarian cancer (n=4), colorectal cancer (n=3), hepato-pancreatico-biliary tract (n=3), malignant epithelioid peritoneal mesothelioma (n=2), and other (n=3). Eleven patients had metachronous PSM. Prior to MDN-PIPAC, seventeen patients had undergone primary tumor resection. The median number of patients had 2 (range: 1-5) lines of systemic chemotherapy before undergoing MDN-PIPAC. Seven patients had previously undergone a median of 2 (range: 1-4) PIPACs with the conventional PIPAC nebulizer in our institutions. The patients’ preoperative baseline data are listed in Table 2.

**Table 2:** Baseline clinical characteristics of the patients with MDN-PIPAC. *Legend: PIPAC = pressurized intraperitoneal aerosol chemotherapy; MDN = multi-directional nebulizer; UGI = upper gastrointestinal; HPB = hepato-biliary tract; ECOG = Eastern Cooperative Oncology Group performance status; PSM = peritoneal surface malignancy*

| Variables | Total population<br>(n = 21) |
| --- | --- |
| Age at first MDN-PIPAC (mean (range) in years) | 62 (38-86) |
| Primary origin (n) |  |
| - Colo-Rectal | 3 |
| - UGI | 6 |
| - HPB | 3 |
| - MPM | 2 |
| - Ovarian | 4 |
| - Other | 3 |
| ECOG (n) |  |
| - 0 | 5 |
| - 1 | 16 |
| Synchronous PSM (n) | 10 |
| Prior primary tumor resection (n) | 17 |
| Prior systemic chemotherapy lines (median (range)) | 2 (1-5) |
| Prior standard PIPAC applications before MDP-PIPAC (median (range)) | 1 (1-4) |
| Ongoing systemic chemotherapy between MDN-PIPAC procedures (n) | 14 |
| The time between primary and metachronous PSM diagnosis (mean (range) in months) | 21 (2-56) |

### 3.6 MDN-PIPAC procedure details, morbidity, and mortality

Abdominal access was achieved in all patients. Thirteen, seven, and one patient underwent one, two, and three MDN-PIPACs, respectively. Two patients received only one cycle of MDN-PIPAC because they were considered candidates for cytoreductive surgery (CRS) and heated intraperitoneal chemotherapy (HIPEC) at the time of staging laparoscopy for MDN-PIPAC.

The median Sugarbaker PCI score and ascites at the first MDN-PIPAC were 25 (1-35) and 200 ml (0-800 ml), respectively. The overall mean (range) operative time was 52.3 (48-78) minutes. The gradual pressure build-up and the subsequent exposure phase of 30 minutes at a capnoperitoneal pressure of 20 mmHg were well tolerated. No patient showed signs of cardio-respiratory impairment from the procedure. No intraoperative surgical or technical problems occurred.

In total, four patients suffered grade I (Dindo-Clavien) complications after a total of 30 MDN-PIPACs (4/30). Three patients experienced persistent nausea and vomiting postoperatively, requiring prolonged intravenous rehydration and antiemetic therapy (without gastric tube) for 48 hours postoperatively. Another patient complained of abdominal pain, which could also be managed after extending intravenous analgesic therapy. No complications grade II-V were observed. The German health system requires a minimum hospitalization of 3 days for PIPAC application in order to be fully reimbursed by the health insurance companies. All patients were discharged on the third postoperative day at home.

## 4.0 DISCUSSION

For more than a decade, a PIPAC nebulizer has been available for off-label clinical use. This technology, with an axially mounted nebulizer unit, has become the standard PIPAC device. Due to its design and performance characteristics, the distribution pattern of the atomized chemotherapy is not homogeneous. Below the opening of the nebulizer unit, approximately 97% by volume is deposited directly onto the underlying peritoneum by inertial impaction [4, 24 - 27]. Recently, other PIPAC nebulizers have become available on the market. However, all nebulizers are based on the same/similar design as the standard technology. Therefore, the problem of local chemotherapy accumulation and inhomogeneous drug distribution pattern in the peritoneal cavity remains unresolved [2], which may affect the oncological efficacy of PIPAC [28]. Therefore, the optimal position of the standard PIPAC nozzle has been investigated in recent years in order to optimize the spatial distribution of chemotherapy [16, 29, 30]. More recently, new experimental PIPAC nozzle technologies have been developed. Rotational intraperitoneal pressure aerosol chemotherapy (RIPAC) uses a modified nebulizer technology similar to the standard PIPAC nebulizer. However, to optimize drug distribution, the nozzle rotates 30° around its vertical axis in the abdominal cavity. In-vivo large animal data indicate that RIPAC leads to a significantly more homogeneous distribution pattern and higher penetration depths of doxorubicin into the tissue compared to the standard PIPAC [31, 32]. Another experimental approach is the use of a preclinical multi-directional nebulizer (MDN). Braet et al. [33] observed in an ex-vivo PIPAC model that the use of their experimental MDN prototype improved the spatial drug distribution of nebulized nanoparticles compared to an SDN. Based on these experimental data, it seems obvious that PIPAC nozzles, which optimize drug distribution, could also be promising in clinical application. For some time now, a CE Class IIa certified MDN has been available for off-label clinical use for MDN-PIPAC [2]. We carried out extensive preclinical testing before introducing this new device into our daily practice.

After the premature discontinuation of a French prospective, randomized phase II study (PIPAC EstoK 01) (https://clinicaltrials.gov/study/NCT04065139) [35], it was vividly discussed whether toxic substances could arise during the application of PIPAC due to the interaction of the chemotherapy solution with the PIPAC nebulizer/syringe/tube system. Therefore, independent toxicological investigations were performed as a first step. For the worst-case scenario with the highest chemotherapy doses, the toxicological investigations showed that no relevant toxicologically questionable substances were observed. The risk assessment was further based on the assumption that three PIPACs are administered consecutively on one day to a patient with a body weight of 10 kg. Even this extreme scenario did not reveal any cumulative toxicological risk for patients. Furthermore, these data indicate that the MDN device could also be used in children with a body weight > 10 kg. It is important to emphasize that our toxicological data are only valid for this type of nozzle. According to our current knowledge, there is no data available for other types of nebulizers.

Our previous and present technical analysis with PIPAC surgeons and scrub nurses in our department showed that the handling of the MDN differs only slightly from that of the standard nebulizer [2]. The MDN unit requires a higher flow rate which can be programmed into the high-pressure injector. However, a major difference is the three additional nozzles built horizontally into the MDN head. Repetitive ex-vivo simulations have shown that the correct mounting of the nozzle head is crucial. The lateral openings of the nebulizers must protrude at least 7 mm beyond the distal end of the trocar for correct atomization.

During the technical risk assessment, concerns were expressed within the PIPAC team that incorrect positioning of the MDN head in the trocar shaft could result in a possible aerosol backflow via the trocar valve into the operating theatre and thus pose a significant risk to healthcare staff. As initial simulations with the ex-vivo PIPAC model did not confirm these fears; we carried out additional schlieren imaging experiments. All of them confirm that even when the MDN head is incorrectly positioned, no CO_2_ is released retrogradely into the environment. Therefore, no backflow of aerosol is assumed based on the CO_2_ visualization.

However, we observed that the MDN can slip into the trocar due to the increased weight of the device. Therefore, we have added a sterile rubber ring to the MDN that is pulled over the nozzle and positioned so that it rests directly on the trocar head, while at the same time achieving the ideal position for the MDN head. Finally, another important safety point is the consistent use of a single-arm holder, which additionally fixes the MDN in a strictly vertical position in the trocar. This measure also prevents the MDN from accidentally tilting sideways during the spraying process, which could affect the spatial distribution pattern of the drug. In order to better monitor this correct position intraoperatively, it would be desirable if the manufacturer could provide a safety mark indicating the ideal position of the MDN head in the trocar. The manufacturer was informed of these points to improve the administrability and safety of MDN-PIPAC.

To further refine the intraoperative MDN-PIPAC safety checklist, based on our previous ex-vivo simulations, we performed MDN-PIPAC in the large animal model immediately followed by CT-peritoneography to visualize the quality of the spatial aerosol distribution. The administration of all three MDN-PIPACs went without any technical or safety problems. The CT-peritoneographies revealed wide aerosol deposition. In particular, areas such as the omental bursa, hepatic interlobar, and deep entero-enteric regions were also affected. Only the ventral parts of the abdominal cavity showed no contrast. However, large gravity-dependent fluid collections, as previously observed with a SDN, did not occur [27].

The clinical application of the elaborated intraoperative MDR-PIPAC safety checklist enabled us to deliver 30 consecutive MDN-PIPACs without intraoperative problems. The postoperative course was characterized by four patients with minor complications (Dindo-Clavien grade I). Based on this initial experience, no clinical difference from SDN-PIPAC was observed at our institution [12].

## 5.0 CONCLUSION

Toxicological data show that the use of the MDN does not generate any leachable substances that could pose a risk to patients. Ex-vivo simulations confirm the importance of correct positioning of the MDN in the trocar and, if possible, in a periumbilical position. Schlieren imaging during ex-vivo MDN-PIPAC simulations showed that even in a worst-case scenario, where the MDN head is incorrectly positioned in the trocar shaft, there is no backflow of CO_2_ via the trocar valve into the environment. Consequently, no backflow of chemotherapy aerosol is assumed. Moreover, our first clinical data on the MDN-PIPAC shows that it is safe to use and causes no problems when the safety checklist is properly adapted to this device. Finally, the perioperative clinical outcome of MDN-PIPAC is comparable to that of our patients previously treated with the SDN. Whether such new devices will have a positive impact on clinical outcomes remains uncertain. Unfortunately, it is unlikely that there will be prospective studies in the near future to clarify this issue. Therefore, we suggest that the type of nebulizer used for PIPAC should also be documented in the ISSPP PIPAC database [35]. Such a large database could provide insight into the role of different nebulizers used in oncological outcomes.

## Data Availability

All data produced in the present study are available upon reasonable request to the authors

## ABBREVIATIONS

AET: Analytical Evaluation Threshold (ISO 10993-18 (2020)
BSA: Body Surface Area calculated according to DuBois
CE: Conformité Européenne
DGAV: Deutsche Gesellschaft für Allgemein- und Viszeralchirurgie
ECOG: Eastern Cooperative Oncology Group
FDA: U.S. Food and Drug Administration
GC-MS: Gas Chromatography coupled with Mass Spectrometry
HSGC-MS: Head-space Gas Chromatography coupled with Mass Spectrometry
IC: Ion Chromatography
ICP-OES: Inductively Coupled Plasma Atomic Emission Spectrometry
LC-MS: Liquid phase chromatography□coupled to a Mass Spectrometer
MDN: Multi-Directional Nebulizer (QuattroJet™, REGER Medizintechnik GmbH, Villigendorf, Germany)
PCI: Peritoneal Carcinomatosis Index according to Sugarbaker
PIPAC: Pressurized IntraPeritoneal Aerosol Chemotherapy
PSM: Peritoneal Surface Malignancies
RIPAC: Rotational intraperitoneal aerosol chemotherapy
SDN: Single-direction nebulizer

## ACKNOWLEDGEMENTS

We are thankful to Aesculap Akademie GmbH, Bochum, Germany for providing the technical equipment to simulate laparoscopic surgery. Especially to Mrs. Anna-Maria Krug, Training Manager at Aesculap Akademie GmbH, Bochum, Germany for her important help. We also thank Mr. Mohsen Faraji and Mr. Burak Kabakci, Technomedics GmbH Illertissen, Germany for providing technical and logistical support for the schlieren experiments in Weimar, Germany. The schlieren flow visualizations were supported by the German Research Foundation (DFG), Grant No. 444059583.

## REFERENCES

1. Di Giorgio A, Macrì A, Ferracci F, Robella M, Visaloco M, De Manzoni G, Sammartino P, Sommariva A, Biacchi D, Roviello F, Pastorino R, Pires Marafon D, Rotolo S, Casella F, Vaira M. 10 Years of Pressurized Intraperitoneal Aerosol Chemotherapy (PIPAC): A Systematic Review and Meta-Analysis. Cancers. 2023; 15(4):1125. 10.3390/cancers15041125

2. Nadiradze G, Horvath P, Sautkin Y, Archid R, Weinreich F-J, Königsrainer A, Reymond MA. Overcoming Drug Resistance by Taking Advantage of Physical Principles: Pressurized Intraperitoneal Aerosol Chemotherapy (PIPAC). Cancers. 2020; 12(1):34. 10.3390/cancers12010034

3. Göhler D, Khosrawipour V, Khosrawipour T, Diaz-Carballo D, Falkenstein TA, Zieren J, Stintz M, Giger-Pabst U. Technical description of the microinjection pump (MIP®) and granulometric characterization of the aerosol applied for pressurized intraperitoneal aerosol chemotherapy (PIPAC). Surg Endosc. 2017 Apr;31(4):1778–1784. doi: 10.1007/s00464-016-5174-5. Epub 2016 Sep 8. PMID: 27631320.

4. Tempfer CB, Rezniczek GA, Ende P, Solass W, Reymond MA. Pressurized Intraperitoneal Aerosol Chemotherapy with Cisplatin and Doxorubicin in Women with Peritoneal Carcinomatosis: A Cohort Study. Anticancer Res. 2015 Dec;35(12):6723–9. doi: 10.1055/s-0035-1560004. PMID: 26637888.

5. Tavernier C, Passot G, Vassal O, Allaouchiche B, Decullier E, Bakrin N, Alyami M, Davigo A, Bonnet JM, Louzier V, Paquet C, Glehen O, Kepenekian V. Pressurized intraperitoneal aerosol chemotherapy (PIPAC) might increase the risk of anastomotic leakage compared to HIPEC: an experimental study. Surg Endosc. 2020 Jul;34(7):2939–2946. doi: 10.1007/s00464-019-07076-3. Epub 2019 Aug 27. PMID: 31456025.

6. Davigo A, Passot G, Vassal O, Bost M, Tavernier C, Decullier E, Bakrin N, Alyami M, Bonnet JM, Louzier V, Paquet C, Allaouchiche B, Glehen O, Kepenekian V. PIPAC versus HIPEC: cisplatin spatial distribution and diffusion in a swine model. Int J Hyperthermia. 2020;37(1):144–150. doi: 10.1080/02656736.2019.1704891. PMID: 32003300.

7. Graversen M, Rouvelas I, Ainsworth AP, Bjarnesen AP, Detlefsen S, Ellebaek SB, Fristrup CW, Liljefors MG, Lundell L, Nilsson M, Pfeiffer P, Tarpgaard LS, Tsekrekos A, Mortensen MB. Feasibility and Safety of Laparoscopic D2 Gastrectomy in Combination with Pressurized Intraperitoneal Aerosol Chemotherapy (PIPAC) in Patients with Gastric Cancer at High Risk of Recurrence-The PIPAC-OPC4 Study. Ann Surg Oncol. 2023 Mar 3. doi: 10.1245/s10434-023-13278-w. Epub ahead of print. PMID: 36867174.

8. Braet H, Andretto V, Mariën R, Yücesan B, van der Vegte S, Haegebaert R, Lollo G, De Smedt SC, Remaut K. The effect of electrostatic high pressure nebulization on the stability, activity and ex vivo distribution of ionic self-assembled nanomedicines. Acta Biomater. 2023 Oct 15;170:318–329. doi: 10.1016/j.actbio.2023.08.027. Epub 2023 Aug 19. PMID: 37598790.

9. Göhler D, Oelschlägel K, Ouaissi M, Giger-Pabst U. Performance of different nebulizers in clinical use for Pressurized Intraperitoneal Aerosol Chemotherapy (PIPAC). medRxiv 2023.03.24.23287646; 10.1101/2023.03.24.23287646

10. Kim G, Tan HL, Sundar R, Lieske B, Chee CE, Ho J, Shabbir A, Babak MV, Ang WH, Goh BC, Yong WP, Wang L, So JBY. PIPAC-OX: A Phase I Study of Oxaliplatin-Based Pressurized Intraperitoneal Aerosol Chemotherapy in Patients with Peritoneal Metastases. Clin Cancer Res. 2021 Apr 1;27(7):1875–1881. doi: 10.1158/1078-0432.CCR-20-2152. Epub 2020 Nov 4. PMID: 33148667.

11. Robella M, De Simone M, Berchialla P, Argenziano M, Borsano A, Ansari S, Abollino O, Ficiarà E, Cinquegrana A, Cavalli R, Vaira M. A Phase I Dose Escalation Study of Oxaliplatin, Cisplatin and Doxorubicin Applied as PIPAC in Patients with Peritoneal Carcinomatosis. Cancers (Basel). 2021 Mar 3;13(5):1060. doi: 10.3390/cancers13051060. PMID: 33802269; PMCID: PMC7958944.

12. Ramos Arias G, Sindayigaya R, Ouaissi M, Buggisch JR, Schmeding M, Giger-Pabst U, Zieren J. Safety and Feasibility of High-Pressure/High-Dose Pressurized Intraperitoneal Aerosol Chemotherapy (HP/HD-PIPAC) for Primary and Metastatic Peritoneal Surface Malignancies. Ann Surg Oncol. 2022 Nov 18. doi: 10.1245/s10434-022-12698-4. Epub ahead of print. PMID: 36400887.

13. Du Bois, D. and Du Bois, E.F. A Formula to Estimate the Approximate Surface Area if Height and Weight Be Known. Archives of Internal Medicine. 1916;17, 863–871. 10.1001/archinte.1916.00080130010002

14. ISO (International Standardisation Organisation). 2020. ISO 10993-18:2020. Biological evaluation of medical devices. Part 18: Chemical characterization of medical device materials within a risk management process

15. ISO (International Standardisation Organisation).ISO 10993-17:2023. Biological evaluation of medical devices. Part 17: Toxicological risk assessment of medical device constituents

16. Giger-Pabst U, Tempfer CB. How to Perform Safe and Technically Optimized Pressurized Intraperitoneal Aerosol Chemotherapy (PIPAC): Experience After a Consecutive Series of 1200 Procedures. Journal of Gastrointestinal Surgery: Official Journal of the Society for Surgery of the Alimentary Tract. 2018 Dec;22(12):2187–2193.

17. Solass W, Giger-Pabst U, Zieren J, Reymond MA. Pressurized intraperitoneal aerosol chemotherapy (PIPAC): occupational health and safety aspects. Ann Surg Oncol. 2013 Oct;20(11):3504–11. doi: 10.1245/s10434-013-3039-x. Epub 2013 Jun 14. PMID: 23765417; PMCID: PMC3764316.

18. Gena AW, Voelker C, Settles GS. Qualitative and quantitative schlieren optical measurement of the human thermal plume. Indoor Air. 2020;30:757–766. 10.1111/ina.12674

19. Alsaad H, Schälte G, Schneeweiß M, Becher L, Pollack M, Gena AW, Schweiker M, Hartmann M, Voelker C, Rossaint R, et al. The Spread of Exhaled Air and Aerosols during Physical Exercise. Journal of Clinical Medicine. 2023; 12(4):1300. 10.3390/jcm12041300

20. Dellweg D, Kerl J, Gena AW, Alsaad H, Voelker C. Exhalation Spreading During Nasal High-Flow Therapy at Different Flow Rates. Crit Care Med. 2021 Jul 1;49(7):e693–e700. doi: 10.1097/CCM.0000000000005009. PMID: 34135285; PMCID: PMC8204857.

21. Meyers MA. Peritoneography: Normal and Pathologic Anatomy. In: Dynamic Radiology of the Abdomen. Springer, New York, NY. 1976; ISBN: 978-1-4757-3957-2

22. Dindo D, Demartines N, Clavien PA. Classification of surgical complications: a new proposal with evaluation in a cohort of 6336 patients and results of a survey. Ann Surg. 2004 Aug;240(2):205–13. doi: 10.1097/01.sla.0000133083.54934.ae. PMID: 15273542; PMCID: PMC1360123.

23. Clavien PA, Barkun J, de Oliveira ML, Vauthey JN, Dindo D, Schulick RD, de Santibañes E, Pekolj J, Slankamenac K, Bassi C, Graf R, Vonlanthen R, Padbury R, Cameron JL, Makuuchi M. The Clavien-Dindo classification of surgical complications: five-year experience. Ann Surg. 2009 Aug;250(2):187–96. doi: 10.1097/SLA.0b013e3181b13ca2. PMID: 19638912.

24. Khosrawipour V, Khosrawipour T, Diaz-Carballo D, Förster E, Zieren J, Giger-Pabst U. Exploring the Spatial Drug Distribution Pattern of Pressurized Intraperitoneal Aerosol Chemotherapy (PIPAC). Ann Surg Oncol. 2016 Apr;23(4):1220–4. doi: 10.1245/s10434-015-4954-9. Epub 2015 Nov 9. PMID: 26553440.

25. Davigo A, Passot G, Vassal O, Bost M, Tavernier C, Decullier E, Bakrin N, Alyami M, Bonnet JM, Louzier V, Paquet C, Allaouchiche B, Glehen O, Kepenekian V. PIPAC versus HIPEC: cisplatin spatial distribution and diffusion in a swine model. Int J Hyperthermia. 2020;37(1):144–150. doi: 10.1080/02656736.2019.1704891. PMID: 32003300.

26. Giger-Pabst U, Bucur P, Roger S, Falkenstein TA, Tabchouri N, Le Pape A, Lerondel S, Demtröder C, Salamé E, Ouaissi M. Comparison of Tissue and Blood Concentrations of Oxaliplatin Administrated by Different Modalities of Intraperitoneal Chemotherapy. Ann Surg Oncol. 2019 Dec;26(13):4445–4451. doi: 10.1245/s10434-019-07695-z. Epub 2019 Aug 9. PMID: 31399820.

27. Bellendorf A, Khosrawipour V, Khosrawipour T, Siebigteroth S, Cohnen J, Diaz-Carballo D, Bockisch A, Zieren J, Giger-Pabst U. Scintigraphic peritoneography reveals a non-uniform 99mTc-Pertechnetat aerosol distribution pattern for Pressurized Intra-Peritoneal Aerosol Chemotherapy (PIPAC) in a swine model. Surg Endosc. 2018 Jan;32(1):166–174. doi: 10.1007/s00464-017-5652-4. Epub 2017 Jun 22. PMID: 28643076.

28. Dedrick RL, Flessner MF. Pharmacokinetic problems in peritoneal drug administration: tissue penetration and surface exposure. J Natl Cancer Inst. 1997 Apr 2;89(7):480–7. doi: 10.1093/jnci/89.7.480. PMID: 9086004.

29. Khosrawipour V, Khosrawipour T, Falkenstein TA, Diaz-Carballo D, Förster E, Osma A, Adamietz IA, Zieren J, Fakhrian K. Evaluating the Effect of Micropump^©^ Position, Internal Pressure and Doxorubicin Dosage on Efficacy of Pressurized Intra-peritoneal Aerosol Chemotherapy (PIPAC) in an Ex Vivo Model. Anticancer Res. 2016 Sep;36(9):4595–600. doi: 10.21873/anticanres.11008. PMID: 27630300.

30. Piao J, Park SJ, Lee H, Kim J, Park S, Lee N, Kim SI, Lee M, Song G, Lee JC, Kim HS; KORIA TRIAL GROUP. Ideal Nozzle Position During Pressurized Intraperitoneal Aerosol Chemotherapy in an Ex Vivo Model. Anticancer Res. 2021 Nov;41(11):5489–5498. doi: 10.21873/anticanres.15362. PMID: 34732419.

31. Mun J, Park SJ, Kim HS. Rotational intraperitoneal pressurized aerosol chemotherapy in a porcine model. Gland Surg. 2021 Mar;10(3):1271–1275. doi: 10.21037/gs-2019-ursoc-11. PMID: 33842275; PMCID: PMC8033043.

32. Park SJ, Lee EJ, Lee HS, Kim J, Park S, Ham J, Mun J, Paik H, Lim H, Seol A, Yim GW, Shim SH, Kang BC, Chang SJ, Lim W, Song G, Kim JW, Lee N, Park JW, Lee JC, Kim HS; KoRIA* trial group. Development of rotational intraperitoneal pressurized aerosol chemotherapy to enhance drug delivery into the peritoneum. Drug Deliv. 2021 Dec;28(1):1179–1187. doi: 10.1080/10717544.2021.1937382. PMID: 34121568; PMCID: PMC8204987.

33. Braet H, Andretto V, Mariën R, Yücesan B, van der Vegte S, Haegebaert R, Lollo G, De Smedt SC, Remaut K. The effect of electrostatic high-pressure nebulization on the stability, activity and ex vivo distribution of ionic self-assembled nanomedicines. Acta Biomater. 2023 Aug 19:S1742-7061(23)00492–0. doi: 10.1016/j.actbio.2023.08.027. Epub ahead of print. PMID: 37598790.

34. Eveno C, Jouvin I, Pocard M. PIPAC EstoK 01: Pressurized IntraPeritoneal Aerosol Chemotherapy with cisplatin and doxorubicin (PIPAC C/D) in gastric peritoneal metastasis: a randomized and multicenter phase II study. Pleura Peritoneum. 2018 Jun 21;3(2):20180116. doi: 10.1515/pp-2018-0116. PMID: 30911659; PMCID: PMC6405009.

35. Mortensen MB, Glehen O, Horvath P, Hübner M, Hyung-Ho K, Königsrainer A, Pocard M, Reymond MA, So J, Fristrup CW. The ISSPP PIPAC database: design, process, access, and first interim analysis. Pleura Peritoneum. 2021 Apr 22;6(3):91–97. doi: 10.1515/pp-2021-0108. PMID: 34676282; PMCID: PMC8482445.

